# Air Pollution Reduction and Mortality Benefit during the COVID-19 Outbreak in China

**DOI:** 10.1101/2020.03.23.20039842

**Authors:** Kai Chen, Meng Wang, Conghong Huang, Patrick L. Kinney, Paul T. Anastas

**Affiliations:** Yale School of Public Health, New Haven, CT, USA; University at Buffalo School of Public Health and Health Professions, Buffalo, NY, USA; Boston University School of Public Health, Boston, MA, USA; Yale School of Forestry and Environmental Studies, New Haven, CT, USA

## Abstract

To control the novel coronavirus disease (COVID-19) outbreak, China undertook stringent traffic restrictions and self-quarantine measures. We herein examine the change in air pollution levels and the potentially avoided cause-specific mortality during this massive population quarantine episode. We found that, due to the quarantine, NO2 dropped by 22.8 µg/m^3^ and 12.9 µg/m^3^ in Wuhan and China, respectively. PM2.5 dropped by 1.4 µg/m^3^ in Wuhan but decreased by 18.9 µg/m^3^ across 367 cities. Our findings show that interventions to contain the COVID-19 outbreak led to air quality improvements that brought health benefits which outnumbered the confirmed deaths due to COVID-19 in China

## LETTER

To control the novel coronavirus disease (COVID-19) outbreak, China undertook stringent traffic restrictions and self-quarantine measures, first in Wuhan and neighboring cities beginning on January 23, 2020, and two days later, in all provinces in China (Figure 1A). The countrywide ban on traffic mobility greatly limited transportation emissions, whereas emissions from residential heating and industry remained steady or slightly declined.^1^ We herein examine the change in air pollution levels and the potentially avoided cause-specific mortality during this massive population quarantine episode.

**Figure 1.**
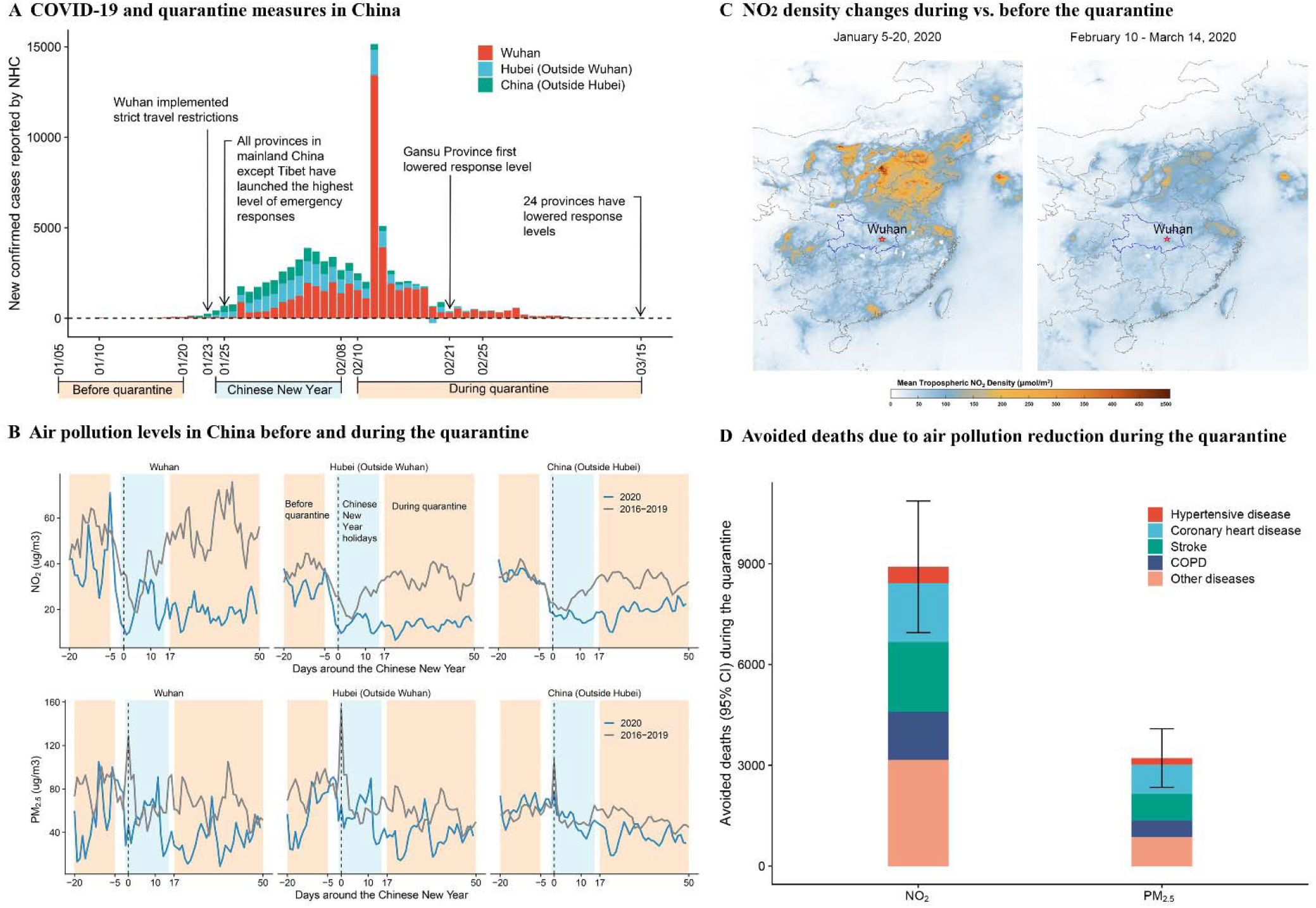
Air pollution levels and avoided cause-specific deaths during the COVID-19 outbreak in China.

As of March 14, 2020, new confirmed COVID-19 cases in China reported by the National Health Commission (NHC) dropped to 20 (4 from Wuhan) (Figure 1A). Most Chinese provinces have lowered the level of emergency responses. We thus defined the “during quarantine” period as February 10 to March 14 and the “before quarantine” period as January 5 to 20. We excluded the Chinese New Year holidays to avoid non-quarantine related air pollution reductions, based on evidence from previous years (Figure 1B). We obtained daily concentrations of nitrogen dioxide (NO_2_) and fine particulate matter (PM_2.5_) in 367 Chinese cities. We then applied a difference-in-difference approach to quantify air pollution changes due to the quarantine. Specifically, we calculated changes in air quality during vs. before the quarantine period in 2020, and compared these with corresponding changes in the same lunar calendar periods in 2016-2019. Details of the material, methods, and numeric results are provided in the Supplementary Appendix.

We found that, due to the quarantine, NO_2_ dropped by 22.8 µg/m^3^ and 12.9 µg/m^3^ in Wuhan and China, respectively. PM_2.5_ dropped by 1.4 µg/m^3^ in Wuhan but decreased by 18.9 µg/m^3^ across 367 cities. The dramatic decline in NO_2_ across China during the quarantine period was also detected by the Copernicus Sentinel-5P satellite using the NO_2_ tropospheric column density (Figure 1C).

We then calculated the avoided cause-specific mortality attributable to these decreases in NO_2_ and PM_2.5_ over China based on the concentration-response functions from a previous study of 272 Chinese cities, and the cause-specific mortality data from the China Health and Family Planning Statistical Yearbook 2018.^2,3^ We estimate that improved air quality during the quarantine period avoided a total of 8911 [95 confidence interval (CI): 6950, 10866] NO_2_-related deaths, 65% of which were from cardiovascular diseases (hypertensive disease, coronary heart disease, and stroke) and chronic obstructive pulmonary disease (COPD), (Figure 1D). Further, we estimate that PM_2.5_ reductions during the quarantine period avoided a total of 3214 (95% CI: 2340, 4087) PM_2.5_-related deaths in China, 73% of which were from cardiovascular diseases and COPD. These numbers should be interpreted with caution due to the potential overlap between PM_2.5_- and NO_2_-related mortality and the disrupted healthcare systems during quarantine to conduct a timely treatment for patients with chronic diseases.

We conclude that interventions to contain the COVID-19 outbreak led to air quality improvements that brought health benefits which outnumbered the confirmed deaths due to COVID-19 in China (3199 deaths as of March 14, 2020).^4^ These findings illustrate the substantial human health benefits related to cardiovascular disease morbidity and mortality that can be achieved when aggressive air pollution control measures are taken to reduce emissions from vehicles, such as through climate mitigation-related traffic restrictions or efforts to accelerate the transition to electric vehicles.

## Data Availability

All data are publicly available.

